# Barriers to surveillance and control of re-emergence of the Chagas disease vector *Triatoma infestans* in Arequipa, Peru

**DOI:** 10.1101/2024.11.27.24318018

**Authors:** Laura D. Tamayo, Valerie A. Paz-Soldán, Carlos E. Condori Pino, Fernando S. Malaga Chavez, Michael Z. Levy, Raquel Gonçalves

## Abstract

Vector control is usually designed as a top-down system with minimum capacity to respond to specificities of epidemiological settings at fine scale or to adjust to routine stressors. Here, we investigated barriers in Chagas disease vector surveillance and control systems in Arequipa, Peru. We conducted in-depth interviews and focus groups with key stakeholders (n=32) at different levels of the health system and community, using process maps to illustrate the workflow for passive and active surveillance. We identified barriers at each step of the process, including systemic, operational, financial, and policy limitations. For passive surveillance, barriers in community participation to report infestations were linked to challenges in capturing the vector and bringing it to a health facility or community health worker. Amongst systemic barriers were related to the use of a data system that did not meet the needs for recording and managing data on vector control activities. At the policy level, the establishment of quotas on the number of houses staff needed to inspect ignores important determinants for infestation and lacks an appropriate sampling design. We discuss the impact of the reported barriers to effective conduction of surveillance and control activities and the initiatives and strategies that have been designed and assessed to bridge these gaps in order to collaboratively design a more resilient health system.

## Introduction

National vector control programs are critical for the prevention and control of vector-borne diseases. Despite well-recognized barriers to existing vector control programs among those managing these programs on the ground, research on the implementation and operational challenges of these remains limited [1]. Time and space for reflection within the system—necessary to identify barriers and develop solutions—are often missing, due to a lack of data, motivation, or an entrenched system that favors traditional strategies over change.

Between 2003 and 2018, a vector control campaign – consisting of an initial preliminary planning phase, an attack phase involving 2 phases of insecticide spraying, and post-spray surveillance– was conducted in the city of Arequipa, Peru, to eliminate the only local insect vector of Chagas disease, *Triatoma infestan*s [2]. Following the campaign, a community-based surveillance system was gradually established, employing both passive and active surveillance methods. In the passive approach, residents are required to capture insects and deliver them to designated health facilities for identification. Once *T. infestans* is confirmed, a vector control specialist inspects the infested house and neighboring homes, subsequently spraying as needed. If *Triatoma infestans* is detected in additional houses, adjacent houses are inspected until the vector is no longer detected [3].

In active surveillance, Vector Control Specialists (VCS) are assigned a quota (20% of households under their supervision) to inspect. Outside of research projects [2,4,5], there is no guidance on *which* households to visit, leaving the selection to the VCS’s discretion. Monthly reports on home inspections are submitted to the Arequipa-Caylloma health network manager.

Our study aimed to examine and depict the barriers within the complex system of triatomine surveillance and control in Arequipa, Peru. We conducted semi-structured interviews with a diverse range of system stakeholders. By triangulating these sources, we moved beyond isolated assessments to co-design more effective and sustainable solutions, enabling the development of more informed and impactful strategies for triatomine surveillance and control in Arequipa.

## Methods

### Study setting

The study was conducted in the Peruvian city of Arequipa (population ~1 million), located in the Andes mountains, south of the capital (Lima). The healthcare system is overseen by two distinct entities: the “Gerencia Regional de Salud” (GERESA), the regulatory body equivalent to a regional ministry of health, and the “Red de Salud Arequipa-Caylloma”, which manages smaller health networks, or “microredes”, to streamline resource allocation and administration within the city. The microredes are clusters of primary care health facilities grouped by geographic proximity, responsible for serving a defined population residing in relative proximity to these facilities. Most health facilities have a “Vector Control Specialist” (VCS) overseeing vector surveillance and control activities in their health network, reporting to both the facility manager and to the GERESA vector control program authority. A few facilities lack a VCS; in these cases, a designated “vector point person” relays vector-related reports to a nearby VCS or directly to the vector control program **authority** in the “Red de Salud Arequipa-Caylloma”.

### Sampling

We used purposive sampling to obtain a range of perspectives regarding different components of the passive and active surveillance system. We interviewed key stakeholders at various levels of the system, including community members, vector control specialists, regional health authorities, and two authorities who have been or are currently part of the vector control unit at the Peruvian Ministry of Health. We conducted interviews and focus groups within each group until thematic saturation was achieved. The semi-structured interviews and focus groups were conducted between November 2021 and March 2024.

### Data Collection and Recruitment

Interviewers used two process maps and flow charts illustrating the passive and active surveillance systems in Arequipa (see Figure 1). These visual aids facilitated discussions with interviewees, encouraging them to specify the challenges at each step of the vector surveillance process. Our objective was explicitly communicated repeatedly before and during the interviews: to establish a collective understanding of systemic barriers.

**Figure 1.**
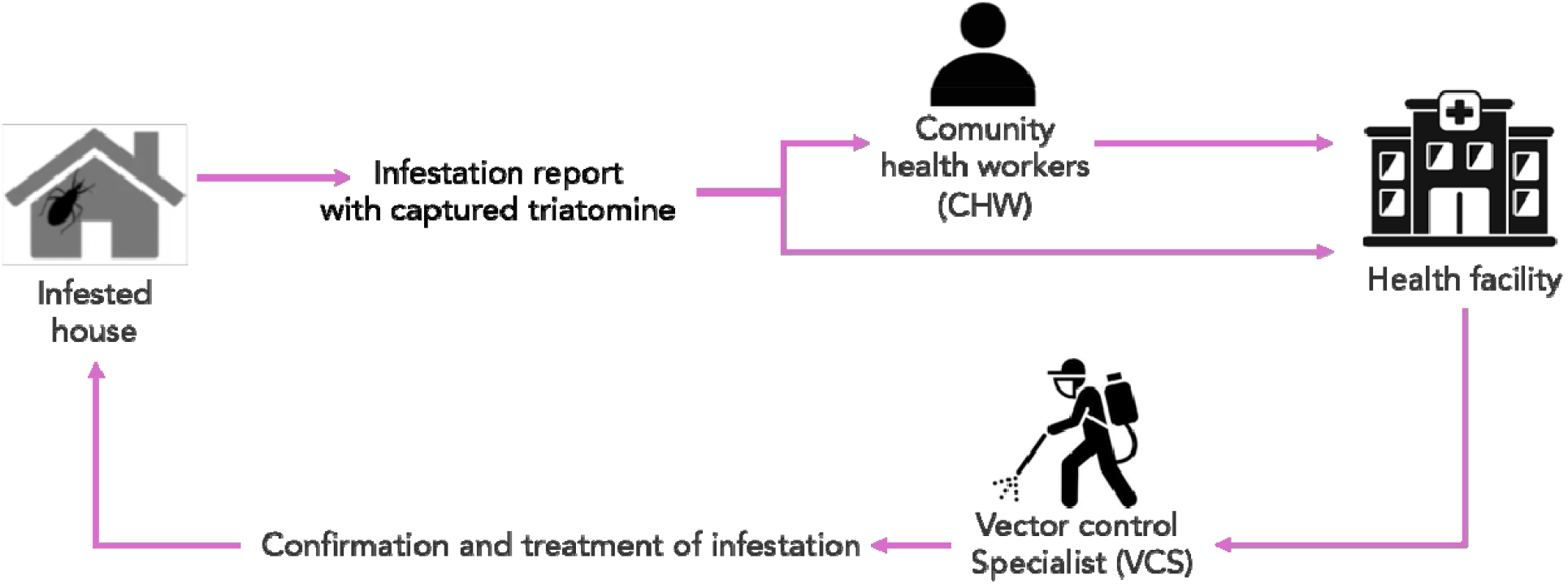
Simple process map of the passive Chagas disease Vector surveillance system in Arequipa, Peru.

Some interviewees, especially Community members, community health workers, and social workers (who oversee the community health workers), had limited awareness of the surveillance system processes. We, therefore, developed a structured interview guide tailored to these participants, focusing on their experiences and challenges within the surveillance system. After interviewing key stakeholders, we held a focus group with vector control specialists to present findings and discuss potential solutions tailored to their unique realities.

### Data Management and Analysis

We audiotaped and transcribed all interviews and the focus group; one respondent requested not to be audiotaped, so we took detailed notes instead. As the interviews progressed, either the interviewer or the respondent filled out blank process maps used to guide discussions on barriers in passive and active surveillance. Based on the initial reading of transcripts, we developed a codebook using both deductive codes (steps of the process maps) and inductive codes (emerging themes). A team of four individuals double-coded all transcripts.

### Ethics statement

The research protocol received approval from the ethical review committees at Universidad Peruana identification number: 833122). Before participation, all individuals provided written consent to take part in the study.

## Results

A total of 32 individuals participated in interviews and focus groups (see Table 1).

**Table 1.** Number of participants recruited and target recruitment by participant role.

| **Role of participant** | **Total Recruited** | **Targeted Recruitment** |
| --- | --- | --- |
| Vector control specialist | 11 | At least one VCS in each “microred” |
| Health authorities | 4 | All surveillance system authorities (and who also manage VCSs) |
| Social workers and Community health workers | 5 | Recruit until saturation related to the Chagas surveillance system reached |
| Community members who reported a home infestation | 10 | All community members who had reported triatomine infestations in their homes through Health facilities or AlertaChirimacha between November 2021 and March 2024 |
| Community members who did <u>not</u> report a home infestation | 2 | Community members who had infestation foci at their homes but who had not reported them |
| **Total** | **32** |  |

We identified multiple barriers at each step of the passive and active surveillance processes, resulting in delays or inadequate response to reports, loss of information, and ultimately, a surveillance system that is not working efficiently. To avoid repetition, we present detailed issues for each step of the passive and active systems in the two figures (Fig. 2 and Fig. 3), while the text focuses on the main themes common to both surveillance systems: 1) systemic challenges and operational barriers, and 2) resource allocation and policy limitations. Additionally, we highlight – and start with – one theme specific to the passive surveillance system: community engagement and reporting challenges.

**Figure 2.**
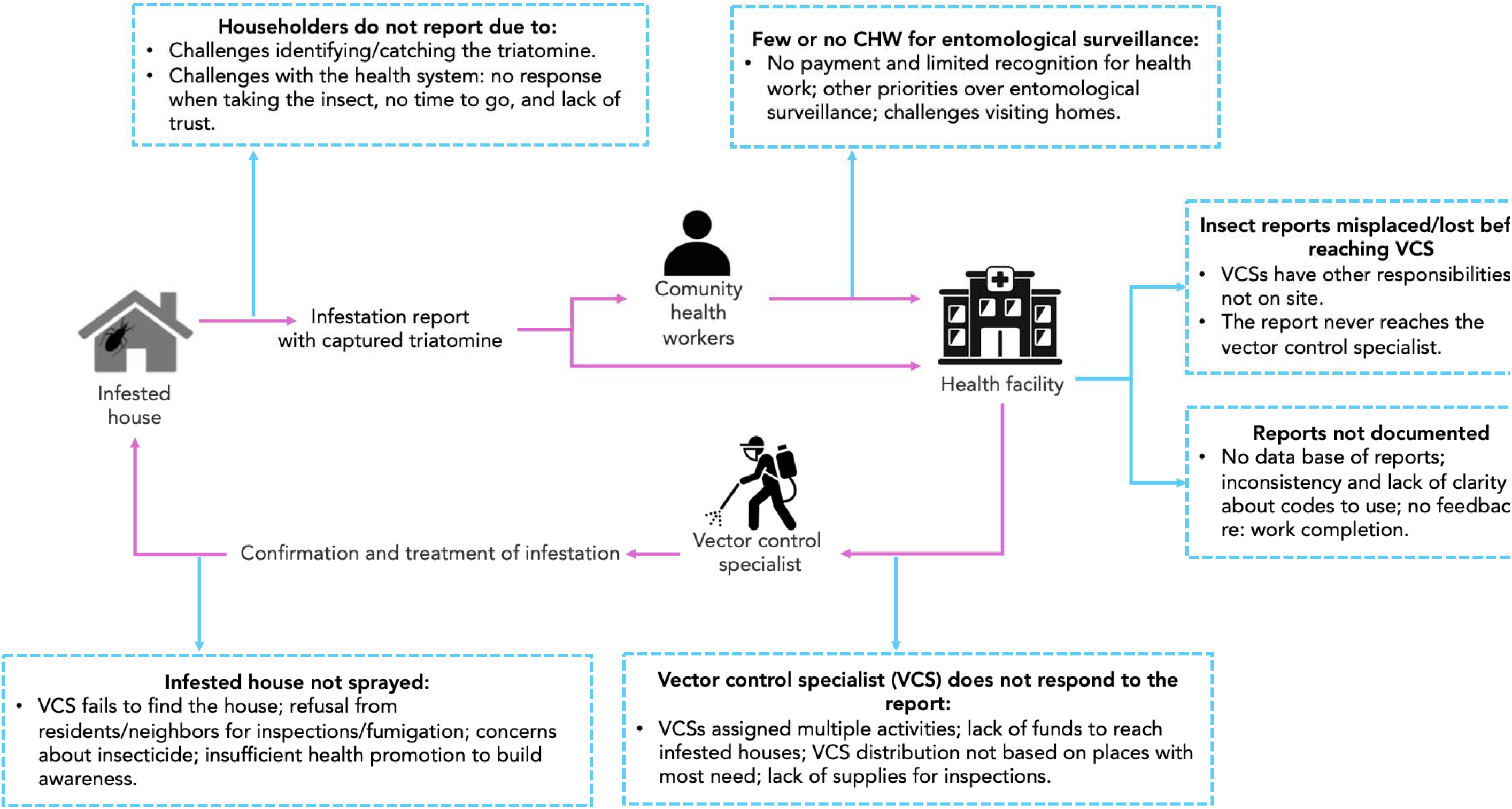
Flowchart of challenges in passive surveillance for triatomine vectors in Arequipa,

**Figure 3.**
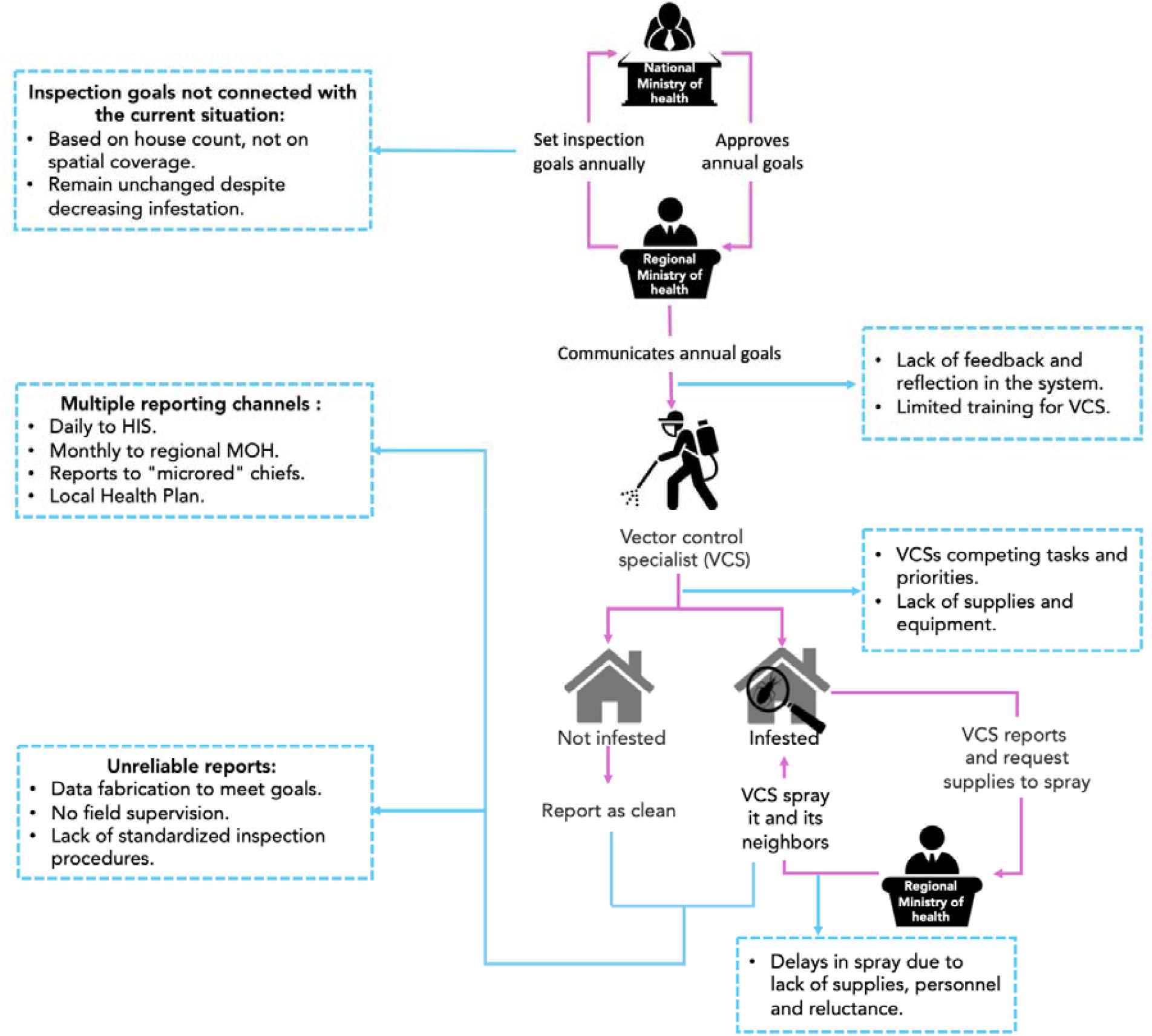
Flowchart of active surveillance for Chagas disease vectors in Arequipa, Peru. Identified barriers are highlighted in light blue boxes.

### Community Engagement and Reporting Challenges

#### Reporting an infestation directly to the health facility

Interviews revealed four main barriers to reporting triatomines to the local VCSs: 1) lack of awareness among community members about identifying the insect and its potential danger; 2) difficulties in capturing it; 3) uncertainty about where or to whom to report; and, 4) receiving conflicting information at health facilities. Currently, community members are expected to capture the insect (usually in a plastic bag or bottle) and bring it to a health facility. The most commonly reported challenge was difficulty in recognizing or capturing the triatomine:

*“*…*I used to see it [*triatomine bug*], but I never thought it was a Chirimacha (*common name for triatomines*), nor did I know that this insect was bad*.*” (Community member)*

*“But they told me, ‘bring it [the Triatomine bug] to me in a bottle’. But that day, as I said, I [thought I] killed it. So I left it. And by the next day, it wasn’t there, it hadn’t died and had gone*.*” (Community member)*

Participants also reported a lack of knowledge about how and where to report infestations. They had difficulties finding time to visit the health center, and prior lack of response from the health system diminished their motivation to do so. One community member who experienced an infestation but did not report it mentioned mistrust in government institutions:

*“Yes, yes. We knew we had to go to report it, but sometimes, always when we go, in any government institution… there is always a delay… and there is a lot of paperwork, a lot of procedures… so you prefer to pay for your own fumigation*” *(Community member)*

#### Reporting an infestation through Community Health Workers (CHWs)

Both CHWs, who can report household infestations, and social workers, who manage the CHWs, noted significant barriers to their work, particularly the absence of financial support and recognition from the health system. This lack of support hinders CHWs’ effectiveness and motivation. As one social worker explained:

> “*[CHWs] do not have money for public transportation, they do not have credentials, their [work] vest is too small and people do not have material, hats, or backpacks…”*

Most community members did not know their CHW or their role. Additionally, most VCSs and those overseeing the vector-borne strategy indicated that CHWs often prioritize incentive-based activities, like a national anemia program, which provides CHWs with a stipend, over vector surveillance. Many stakeholders, including social workers and VCSs, mentioned that CHWs are often not well-known within the health system, partly due to high staff turnover within the system, and are not fully integrated into the vector surveillance system. One VCS described:

> *“If you ask me to bring you a [CHW], I don’t even know their address… I only know them when they come to a health promotion meeting, and later I might see them again when it’s Chagas week in November when they tell us we have to come to the regional government with community health workers. That day, I go to the [*the social worker*], and say, ‘please, assign me some [CHWs], and I’ll bring them’”. (Vector control specialist)*

### Systemic challenges and operational barriers

#### Delays and mismanagement in reporting

Community members reported significant delays in responses to infestation reports, often requiring multiple visits to the health facility before action was initiated. When VCSs were absent, other health staff mishandled reports by misidentifying the triatomine, dismissing the report, or failing to relay the information to the VCS. In some cases, residents were asked to return later or the next day to submit the insect.

> *“*… *if I am not there, sometimes they send it [a triatomine] to the laboratory, other times it stays in the admission area and sometimes it gets lost*… *perhaps I am told they brought one, but I don’t remember because the next day they cleaned, saw a bag with an insect and maybe threw it away*.*” (Vector control specialist)*

##### Interviewer

> *When the community member arrives with a chirimacha, and it is received in the admission area, do they take it to you or does it go to the laboratory? Is there a sheet or documentation of everything that has happened [hand off of the triatomine]?*

##### Vector control specialist

> *No*…, *no, there is no record, we should do better about that*.

#### Resource constraints

Vector control specialists are often tasked with other activities, such as patient admission, logistics, triage, and water and sanitation quality control, as well as responding to other zoonotic disease reports. VCSs reported finding it difficult to refuse these tasks since they are assigned by their direct supervisor. They also lack resources, including equipment, supplies, and transportation funds, to carry out inspections or spraying.

#### Resident reluctance and integrity concerns

In addition to challenges reaching the field for surveillance, VCSs encounter barriers during inspections, such as residents’ fear and reluctance to let them in their homes, due to growing insecurity. This occurs despite prior coordination with the health promotion unit, which works to raise community awareness about the importance of surveillance and allowing VCSs’ entry. One VCS described:

> *“Yes, insecurity, insecurity is strong*… *that is one of the biggest obstacles, right now [*…*]. That is the biggest problem, they are reluctant, right? They are afraid. People live now with a lot of fear, a lot of fear. They don’t open the door easily or if they answer, they answer from the roof or the window*.*” (Vector control specialist)*

Neighbors of an infested index house sometimes refuse inspections and treatment, complicating efforts to control infestation foci. One VCS also observed reluctance to allow spraying due to previous negative experiences:

> “*Another thing that sometimes happens is that they [community members] are afraid, they are not very confident [of the insecticide]*… *[We are asked:] ‘What are we going to fumigate and with what?’ Why? ‘Because the previous neighbor or the previous fumigations had*… *what is it called? Hypersensitivity to insecticide…’ and no, they don’t want [to fumigate]*.” *(Vector control specialist)*

Both VCSs and health authorities raised concerns about the quality and consistency of house inspections – and even if they are being conducted at all. They noted some VCSs no longer perform thorough household inspections. Some only ask residents about triatomines in their home, others enter only parts of the home, whereas others might perform very quick, superficial checks. Authorities also reported instances in which some VCSs falsified data to meet established goals:

> “*[One vector control specialist] would come with his little sheet of paper, because we have to have our tangible proof [of inspected houses]. At some point, they told us that homeowners had to sign a paper. But, how do I believe him? Because I have found several papers from previous vector control specialists, I don’t know how they did it, but the code [house identifier] did not match*.. *I went there myself once because as I am responsible: [the codes] did not match. They didn’t match at all. That’s my problem. So I told [the VCS]: ‘If you don’t do it right, I will always check your little sheet of paper [documentation of inspected house] I’ll do random checks’. And some may respond, ‘Yes, yes you are right, excuse me’. [I tell them]: ‘Don’t fool me, because I know my area’. So I tell you, maybe they made up data to cover the quotas that they had?*… *I don’t know*.” (Lead vector control specialist)

#### Competing priorities and quality related to the work of Vector Control Specialists

VCSs report to both the health facility manager and the vector control authority, juggling competing tasks such as sanitation, water quality control, health facility assistance, and vector surveillance. As a result, VCSs feel pressured to complete tasks quickly, often with minimal effort, to meet their goals.

> *“Many times the work is based on the convenience of the health staff and I would imagine that the inspection, if they do it at all, is going to be as quick as possible*…*” (Health Authority)*

The frequent turnover of facility managers aggravates this problem by assigning non-surveillance tasks to the VCSs. A health authority noted that each new manager has different priorities for VCSs, impacting their ability to focus on active surveillance. As a result, the health authority often has to “fight” to ensure VCSs fulfill their surveillance duties:

##### Interviewer

> *And the change of [health facility] managers, how much does it affect surveillance?*

##### Health authority

> *“It affects because a new manager comes in with a new idea. ‘Ah, right! You, for example, you are not going to do that, you are going to do other things, because I need here in this service because here I have no one to do it, so you are going to do it*.*’ ‘But, doctor, I have my [surveillance] activities’. ‘Just leave those activities!’ There is no importance given to the [surveillance] program on behalf of the managers*.*”*

### Resource allocation and policy limitations

#### Budget allocation determination at the national and regional levels

The Ministry of Economy and Finance distributes the budget for zoonotic diseases across government levels. Approximately 22% remains with the National Health institutions (MINSA, CDC, INS) 74.8% is allocated to regional governments, and 3.2% to local governments (data from 2022) [6]. To secure funding, the Regional Ministry of Health (GERESA) must plan and budget for active surveillance activities in line with the technical standards set by the National Ministry of Health. However, regional health authorities reported that the allocated budget is usually less than requested, forcing them to re-adjust their plans. The Peruvian health system incorporates ‘pay for performance’, in which budget allocation is based on the completion of established activities or specified goals, but the budget is often reduced even if goals are purportedly met. One health authority describes:

##### Interviewer

> *So what they give you depends on your results?*

##### Health authority

*No, no. What they call pay for performance…I think that was the original idea: if you give me health results, then I increase your budget, as a ‘reward’ so that you can continue to carry out your activities and move forward. But no, you meet the goals, and they don’t give you anything, they reduce it [the budget]. Then they say: ‘but you are not complying*.*’ Of course, I am not complying because I have no means to comply. ‘No, but you must comply in order to…’ How am I going to comply?! If I have no money to be able to comply, how do you want me to comply? [You want me to] make it up just to look good?*

On the other hand, regional governments allocate their funds based on their priorities. While part of the budget is reserved for nationally prioritized health strategies, currently anemia and tuberculosis, the remaining funds are distributed at the discretion of regional governments. Local health authorities for vector-borne and zoonotic diseases noted that Chagas and other zoonotic diseases are often not prioritized, leading to competition between health units to maintain or increase their budgets. One local health authority elaborated further:

> *“It would be different if, say, we had a conversation, raised awareness, or did some advocacy with the regional government so that they could increase our budget. We don’t want billions, but we do want to have enough to sustain the existing achievements. For Chagas, we have great achievements, but these can’t be sustained because there is not enough funding…”*

Another health authority added:

> *“The regional government distributes [money] according to its priorities. That is why the management coordinators, for example, go and fight, right? saying: ‘This is my budget and nobody touches it*… *because I need it for this, for that, because people with TB are going to die because pregnant women are going to die*.*’ So the regional government says, ‘nobody touches those budgets’ and if there is extra, they give it to the other [diseases]…”*

#### Surveillance Goals and Metrics: Lack of Clarity Guiding Decision-Making and Planning

According to national guidelines [7], 20% of homes at risk for triatomine infestation must be inspected annually. This target is set without considering evidence-based definitions of ‘risk area’, nor the amount of inspections conducted previously, nor the spatial coverage of inspections. Below are some excerpts from a local health authority and a VCS, depicting this lack of clarity and its impact on motivation to perform work thoroughly:

> *“The [surveillance] goals remain the same; what [should] change is where the inspection is carried out. But the vector control specialist inspects in the same places because it’s easier*…*” (Health Authority)*
>
> *[…] “If it is the same area, you go year after year, year after year, to a house. They tell you you have to enter [the houses], to inspect and all that. But [if you’ve been to the house before] and you know that you have never found it* [the triatomine], *you do it a little bit faster…” (vector control specialist)*

VCSs reported that inspections take anywhere from 10 to 30 minutes, indicating significant variation in thoroughness. Additionally, a health authority noted that the national guidelines for triatomine surveillance have not been updated in decades:

> *In Arequipa we work based on our experience. The [National] Chagas standard is outdated…, it is from the year 1998 and it is in ‘the process of being updated’, but no one knows where it’s really at. For many years we have been asking for it… it has to be done by MINSA [the Peruvian Ministry of Health]”. (Health Authority)*

Regional authorities also explained that the electronic system used to report programmed activities is incomplete, so they also rely on a parallel paper-based system. This dual reporting generates further complications, as the Ministry of Health and the Ministry of Economy and Finance base budgetary decisions on the electronic system, which fails to capture the full extent of activities conducted, making it harder to secure adequate funding. Additionally, sometimes VCS visit homes for follow-up, but do not find the resident at home, there is no way to document, or receive reimbursement, for this expense. One local health authority explains in detail:

> *“The State has implemented electronic systems for me to receive money and to report what I am spending. But those systems have their flaws. A lot of the information doesn’t get through. Lima tells me: ‘Hey, you are not working’, but I am working: look at my reports, look at my documents! [They respond:] ‘But I don’t have it here in the system’ because the failure starts here and it gets lost along the way… Lima enters into the [electronic] system… and says: ‘Let’s see - are you producing? No! You are not producing anything! So, what money should I give you, if you don’t need it?’ So that’s where the budget starts getting lost*.*” (Health Authority)*

#### Inconsistencies and Limitations in the Reporting System

Our respondents also discussed barriers in reporting vector surveillance data due to a lack of standardized reporting procedures, including missing reporting codes in the Health Information System (HIS), data that HIS requests but does not report out (see first excerpt below), and the lack of a way to document visits to houses that denied entry to the VCS or were unavailable at the moment of the visit (second excerpt). Additionally, reporting practices among VCSs vary widely: some only report monthly to the vector-borne strategy authority, others only report through the HIS, and a few also report their activities in the local health plan. Additionally, one participant mentioned reporting to the local municipality.

> “*The HIS problem*… *Do you know what the problem is? We put information in HIS, but it is not reported in the report. In other words, the electronic system takes in the information but does not report it. So, at the end of the month, I want to know how much was done, [but] nothing comes out! Why am I going to report it, if nothing comes out?*” (Vector Control Specialist)

Regarding the inability to report “unsuccessful” house visits (where they were unable to speak to the resident for different reasons), one VCS explains.:

> *In other words, in our document, we are only asked for the number of homes inspected during the month of April, for example, 30 [houses]. But a house that did not let you go in, no, that doesn’t count, you have to go again. (Vector control specialist)*

## Discussion

This study identified critical gaps and barriers in the processes of the vector surveillance system for Chagas disease in Arequipa. At each step, challenges emerged that hindered the system’s efficiency, including fragmented information flow, inadequate documentation and follow-up, insufficient health personnel for responses, and a lack of “reflection” in a complex and dynamic system that did not incorporate new goals or strategies. Collectively, these barriers significantly diminish the system’s potential to control the spread of triatomines effectively. Our findings are consistent with those from other regions of the Southern Cone [8–11], where the efficacy of triatomine control programs is compromised by a lack of sustained political commitment, decentralization of disease control programs, a reduction in trained personnel responsible for vector control operations, and the expansion of dengue and other outbreaks which divert resources away from Chagas disease vector control [12–14].

Active surveillance for vector control in Arequipa operates on a quota system (“metas”), requiring personnel to inspect a fixed number of houses for triatomines. Quotas, and pay-for-performance schemes [15,16], are strategies imported from high income countries [17]. These have led to Catch-22’s: personnel must meet quotas to receive funding, but need funding to pay for transport and other costs necessary to meet the quotas. While the quotas come from the leadership of the vector control program, the day to day activities of the vector control specialists are under the supervision of health center managers. Vector control specialists must negotiate with their immediate managers for the resources and time they need to conduct household inspections. Managers rarely support active inspections, and often assign vector control specialists to unrelated tasks at the health center. Coordination amongst government institutions, and the tensions it generates, is a consequence of the decentralization in health [18] - an imposed reform that shifted authorities to regional, district, and municipal levels [19]. Experiences across Latin America report multiple negative impacts of decentralization, including the absence of local expertise [20,21], limited resources and organization [22], and lack of local political leadership to sustain the programs [22]. In Argentina, for example, it took 10 years following decentralization for regional programs to achieve the same coverage of Indoors Residual Spraying they had achieved before the reform [23]. In Brazil, decentralization led to unsystematic and sporadic data collection, leading to the loss of valuable epidemiological information [24]. In Minas Gerais State, Brazil, domestic Chagas disease vector infestation persists due to the inefficiency of municipal programs - some of which have not conducted any control activity since decentralization [22].

Further, the quotas are simply a *number* of houses--they are agnostic in terms of *which* houses are to be inspected. Facing the additional constraint of lack of transportation funds, and limited time and support from direct superiors for active surveillance, most vector control specialists, if they conduct inspections at all, do so through convenience sampling--visiting households near the health centers, or occasionally near water treatment plants or other installations they visit for other purposes. The quota system must be de-implemented to make space for more rational and effective approaches, especially those that can make use of the wealth of spatial information, historical spray data, and other known correlates of infestation risk [25] to guide active surveillance efficiently. Neglecting such information about population susceptibility to vectors, and providing insufficient funds for vector control specialists to reach at-risk households, contributes to reinforce inequalities in health coverage and delivery.

Unnecessary barriers and inefficiencies similarly plague the passive surveillance system. The requirement for community members to capture and bring a triatomine to a community health worker or a health center is impractical and discourages participation. Community members can be reticent, or unable, to capture the insects. They may lack time or motivation to visit a health center or may feel uncertain that their efforts will lead to an effective response. Very few community health workers are ‘active’, and those often spend more time and energy on better-funded programs. The health posts do not have a good system for receiving triatomines. Insects brought in by community members are frequently lost, mishandled, or forgotten. Although passive surveillance has been reported as a sensitive and cost-effective method for early detection of infestations [26,27], it relies on key elements, such as a close relationship between affected communities and researchers and/or surveillance staff [26]; a quick response to infestation reports [28], as well as a sustained community motivation to participate in surveillance activities [29]. In endemic areas where these elements are missing, community participation in passive surveillance presents similar barriers to the ones described in Arequipa [22,30]. Addressing these challenges requires revising passive surveillance protocols to simplify reporting, foster trust, and strengthen the connection between communities and Health systems.

Data flow modernization is critical to the effective functioning of the surveillance and control system, being a key element to deliver health services [31], providing reliable data to support informed decisions in health [32], and improving health system response to stressors and shocks [33]. The Health Information System (HIS) in place to record triatomine surveillance and control activities in Arequipa was originally developed to manage primary healthcare data and related administrative activities [34]. It is rigid and structured and does not address the needs of the triatomine surveillance system, neither active nor passive, resulting in barriers to data recording, management, flow, and analysis. Participants report that entomological inspections and infestation data are often incomplete, geospatial information critical for planning interventions is missing, and data flows only one way, preventing analysis to adjust interventions as needed. The current data system does not allow an assessment of the impact of surveillance and control activities. Some activities are conducted but not registered, or staff might be discouraged to conduct activities that will not be visible in the system [30].

New strategies for triatomine vector surveillance and control have recently been tested in Arequipa. These show promise to build resilience for routine stressors and unexpected shocks by strengthening the system’s adaptive response and its capacity for self-regulation [35]. In a cluster randomized trial, reframing the surveillance system from a militarized, top-down system to one patterned after the immune system [36], significantly increased the system’s ability to detect and eliminate infested houses compared to a conventional approach. A new internet-based community reporting system [3], not only ensured the continuous delivery of services under an unexpected shock (the SARS-Covid pandemic) but also increased the system’s ability to detect infestations. While these strategies have proven successful in Arequipa, they clash with existing rules and policies, complicating their integration into the broader health system [33,37].

This evaluation was conducted in a specific city within a particular context; the results are not meant to be generalized across Peru, or beyond. However, the findings highlighted barriers that, although specific to the entomological surveillance system, may similarly affect other areas of the Peruvian health system [19], as well as vector surveillance systems elsewhere [39]. In addition, we could not interview health authorities at the national level, which leaves a gap in understanding the higher-level policy decisions that shape the implementation of surveillance systems. Furthermore, the limited number of social and community health workers available for interviews (due to their scarcity) constrained our ability to explore their perspectives and experiences comprehensively.

Vector-borne diseases are dynamic systems, requiring constant adaptation to changes in vectors, pathogens, environments, and societal conditions. To remain effective, surveillance and control systems must integrate new strategies and interventions that can keep pace with the constant changes in these dynamic systems. When a newly implemented strategy succeeds, changes the epidemiological scenario, which in turn will require a constant cycle of evaluation and innovation [40]. Achieving such a system requires participatory approaches that actively engage affected communities, health authorities, and other government sectors [33]. This collaborative framework ensures the system can operate flexibly and adaptively, functioning with multiple interconnected components to address diverse needs effectively. At the policy level, integrating vector surveillance and control with other government sectors, such as urban planning, education, and environmental management, is essential [33,41]. Vulnerability to Chagas disease vectors is driven by underlying social determinants [42–44], and addressing these factors through coordinated policies can alleviate the disease burden on affected populations and health systems alike. Building resilient systems capable of adapting to routine stressors and unexpected shocks requires commitment at all levels— local, regional, and national—and a shift from reactive to proactive, integrated approaches to health system strengthening.

## Data Availability

All data produced in the present study are available upon reasonable request to the authors

